# Age-dependent association between SARS-CoV-2 cases reported by passive surveillance and viral load in wastewater

**DOI:** 10.1101/2021.04.30.21256366

**Authors:** Ryosuke Omori, Fuminari Miura, Masaaki Kitajima

## Abstract

The true number of individuals infected with severe acute respiratory syndrome coronavirus 2 (SARS-CoV-2) is difficult to estimate using a case-reporting system (i.e., passive surveillance) alone because of asymptomatic infection. While wastewater-based epidemiology has been implemented as an alternative/additional monitoring tool to reduce reporting bias, the relationship between passive and wastewater surveillance data has yet to be explicitly examined. Since there is strong age dependency in the symptomatic ratio of SARS-CoV-2 infections, this study aimed to estimate i) an age-dependent association between the number of reported cases and the viral load in wastewater and ii) the time lag between those time series. The viral load in wastewater was modeled as a combination of contributions from different age groups’ virus shedding, incorporating the delay, and fitted with daily case count data collected from the Massachusetts Department of Public Health and SARS-CoV-2 RNA concentrations in wastewater collected by the Massachusetts Water Resources Authority. The estimated lag between the time series of viral loads in wastewater and of reported cases was 10.8 days (95% confidence interval =[10.2, 11.6]) for wastewater treatment plant’s northern area and 8.8 days [8.4, 9.1] for southern area. The estimated contribution rate of a reported case to the viral load in wastewater in the 0–19 yr age group was 0.38 [0.35, 0.41] for northern area and 0.40 [0.37, 0.43] for southern area, that in the 80+ yr age group was 0.67 [0.65, 0.69] for northern area and 0.51 [0.49, 0.52] for southern area. The estimated lag between those time series suggested the predictability of reported cases ten days later using viral loads in wastewater. The contribution of a reported case in passive surveillance to the viral load in wastewater differed by age, suggesting a large variation in viral shedding kinetics among age groups.

## 1. Introduction

Following the emergence of severe acute respiratory syndrome coronavirus 2 (SARS-CoV-2) in 2019 (1), the epidemic has resulted in a large number of deaths in many countries. The development of effective pharmaceutical interventions for eradication is ongoing, and non-pharmaceutical interventions (i.e., restriction of movement) are still required to reduce the risk of mortality. For the latter to be effective, an understanding of the current state of the epidemic is needed. In infectious disease epidemiology, the time series of the number of confirmed cases is monitored (so-called passive surveillance) and used to understand the current state of an epidemic. However, in the context of the COVID-19 pandemic, such data have large biases because the testing frequency and propensity are not constant over time, so the sampling process is non-random (2). For instance, the number of confirmed cases strongly depends on the number of people who recognize their infection or suspicious contact with infected individuals. SARS-CoV-2 is known to cause asymptomatic infections and the rate of symptomatic infection depends on age (3, 4). These characteristics make it more difficult to estimate the true number of incidences from the number of confirmed cases, requiring an alternative approach to identifying these potentially unreported transmissions.

In this context, WBE has drawn attention from scientists (5) and public health authorities (6, 7). Even during the initial phase of the epidemic, SARS-CoV-2 RNA detection in wastewater was reported in various countries such as Australia (8), Japan (9), the United States (10), and many countries in Europe (11, 12). The Netherlands has already implemented a nationwide wastewater monitoring system (13), and more than 40 states in the United States have utilized WBE (14, 15). While this has potential as an early warning system, considerable challenges remain (16). For instance, although several studies have conducted lagged correlation analysis (17-19), there exists no explicit modeling method that can relate the observed viral load in wastewater to the number of infected individuals.

To capture trends in ongoing transmissions from the viral load in wastewater, it is essential to understand their quantitative relationship. While the detection rate by passive surveillance depends strongly on age, resulting in a bias in the number of confirmed cases, in principle the viral load in wastewater reflects actual infection levels in communities as long as the wastewater monitoring methods (i.e., condition of influent samples, timing of wastewater sampling, virus detection protocols, etc.) are consistent over time. Therefore, a comparison between passive surveillance and wastewater surveillance could be expected to quantitatively measure age-dependent biases in passive surveillance.

The present study aimed to estimate the age-dependent association between the number of reported cases in passive surveillance and the viral load in wastewater to compare passive and wastewater surveillance methods. We constructed a statistical model describing the association between incidence per age group and the viral load in wastewater, then estimated the lag of detection timing between passive surveillance and wastewater surveillance and age dependency in the contribution rate of incidences to the viral load in wastewater.

## 2. Methods

### 2.1. Data collection

Daily case count data were collected from situation reports produced by the Massachusetts Department of Public Health (20). The definition of a “confirmed” case was a patient who met confirmatory laboratory criteria (i.e., detection of SARS-CoV-2 RNA in a clinical or autopsy specimen using a molecular amplification test) (21). Since age-specific data were available only until August 11, 2020, we analyzed the case series from March 22 through that date.

For wastewater, we used data from the Massachusetts Water Resources Authority, in which SARS-CoV-2 RNA concentrations were measured by Biobot Analytics (22). The wastewater treatment facility used has two major influent streams (“northern” and “southern”), and wastewater samplings were conducted 3–7 times per week for each influent stream. The concentration of SARS-CoV-2 was normalized to the concentration of a human fecal indicator (PMMoV) in order to compensate for variations in fecal load in wastewater, which can be affected by factors such as fecal excretion patterns and dilution. Details of the sampling and molecular methods were described in a previous study (19).

### 2.2. Model

To understand the heterogeneity in the contribution of incidence to the viral load in wastewater by age, age-group-specific incidences were compared to the viral load in wastewater. Assuming that the lag between the detection timing from wastewater and the reporting by passive surveillance is similar between age groups, the viral load in wastewater at time *t, w*(*t*), can be written as:

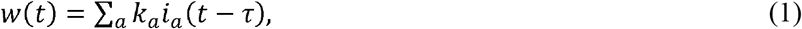

where τ denotes the lag between the detection timing from wastewater and reporting by passive surveillance, *i*_*a*_ denotes the incidence in age group *a*, and *k*_a_ denotes the contribution rate of an incidence in age group *a* to the viral load in wastewater. This model was fitted to the data for viral load in wastewater *w*_*data*_(*t*) by maximizing the likelihood function considering the Poisson sampling process, *L*:

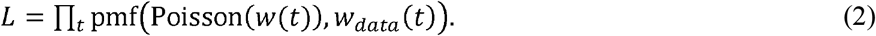

We then calculated profile likelihood-based confidence intervals for each parameter. In the model, *i*_a_(*t*) was defined as the change in continuous time, but the sampling times of real data were discrete. To obtain this variable in continuous time, we determined the two-week moving average of the reported cases from passive surveillance by cubic spline interpolation.

To understand the heterogeneity in the contribution of age-group-specific reported cases, we conducted a multiple regression analysis between the number of reported cases by passive surveillance and the viral load in wastewater. The coefficients in the model, *k*_a_s, were compared to evaluate the contribution of each age group. The relative evaluation of coefficients in multiple regression is biased by multicollinearity if the explanatory variables (in our study, the number of reported cases per age group) are strongly correlated. To reduce this bias, we calculated the Pearson correlation coefficients between pairs of explanatory variables in the model (*i*_*a*_ in Equation (1)). If a correlation coefficient > 0.7 was found, one of two explanatory variables was excluded until all correlation coefficients were < 0.7. All of the above computations were implemented in Mathematica ver. 12.0.0.0.

## 3. Results and discussion

### 3.1. Multicollinearity in the regression model of viral load in wastewater

Strong correlations in the number of reported cases by passive surveillance between age groups were found (Table 1), which expected a large bias in the multiple regression analysis. As shown in Fig. 1, the time-series trends in the number of reported cases between the close age groups were similar. This similarity can be considered to induce multicollinearity in the regression model. The time series of reported cases by passive surveillance in age groups 0–19 years and 80+ years were chosen as the explanatory variables to satisfy the criteria. In the selected model, all correlation coefficients between explanatory variables were < 0.7.

**Table 1.** Pearson correlation coefficients of the time series of reported cases by passive surveillance between different age group pairs.

|  |  | Age group (yr) |  |  |  |  |  |  |
| --- | --- | --- | --- | --- | --- | --- | --- | --- |
|  |  | 20–29 | 30–39 | 40–49 | 50–59 | 60–69 | 70–79 | 80+ |
| Age group | 0–19 | 0.78 | 0.71 | 0.67 | 0.63 | 0.60 | 0.45 | 0.44 |
|  | 20–29 |  | 0.96 | 0.94 | 0.94 | 0.91 | 0.77 | 0.70 |
|  | 30–39 |  |  | 0.99 | 0.97 | 0.97 | 0.87 | 0.82 |
|  | 40–49 |  |  |  | 0.98 | 0.98 | 0.89 | 0.84 |
|  | 50–59 |  |  |  |  | 0.98 | 0.89 | 0.83 |
|  | 60–69 |  |  |  |  |  | 0.93 | 0.88 |
|  | 70–79 |  |  |  |  |  |  | 0.97 |

**Fig. 1.**
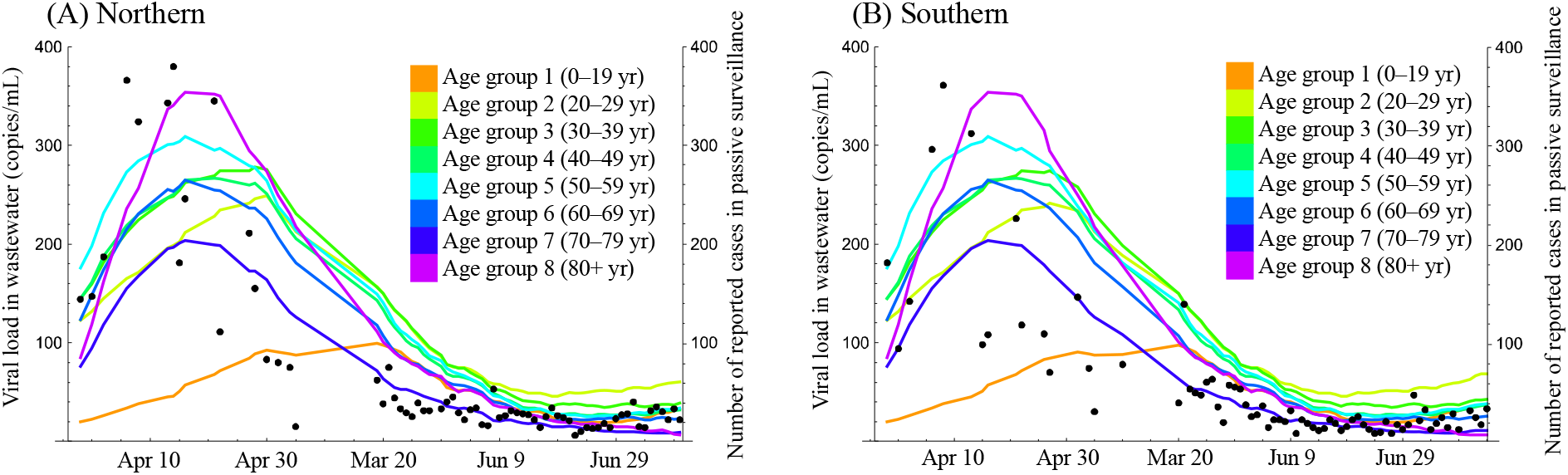
Comparison between viral load in wastewater and the number of reported cases per age group in passive surveillance. Dots show viral load in wastewater sampled in the treatment plant’s (A) northern and (B) southern area. Solid lines show the two-week moving average of the number of reported cases per age group in passive surveillance, with color representing age group.

### 3.2. Model fittings

We used the model described in Equation (1) to estimate the lag between the detection timing from wastewater and the reporting from passive surveillance, τ, and the contribution rate of an incidence in age group *a* to the viral load in wastewater, *k*_a_. The maximum likelihood estimates of τ were 10.8 days (95% confidence interval [95%CI] = [10.2, 11.6]) for the northern area and 8.8 days (95%CI = [8.4, 9.1]) for the southern. The maximum likelihood estimates of *k*_a_ for the age group 0–19 yr were 0.38 (95%CI = [0.35, 0.41]) for the northern area and 0.40 [95%CI = [0.37, 0.43]) for the southern. The maximum likelihood estimates of *k*_a_ for the age group 80+ yr were 0.67 (95%CI = [0.65, 0.69]) for the northern area and 0.51 (95%CI = [0.49, 0.52]) for the southern. The model’s estimated parameters reasonably explained the data for viral load in wastewater sampled in both the northern and southern areas (Fig. 2).

**Fig. 2.**
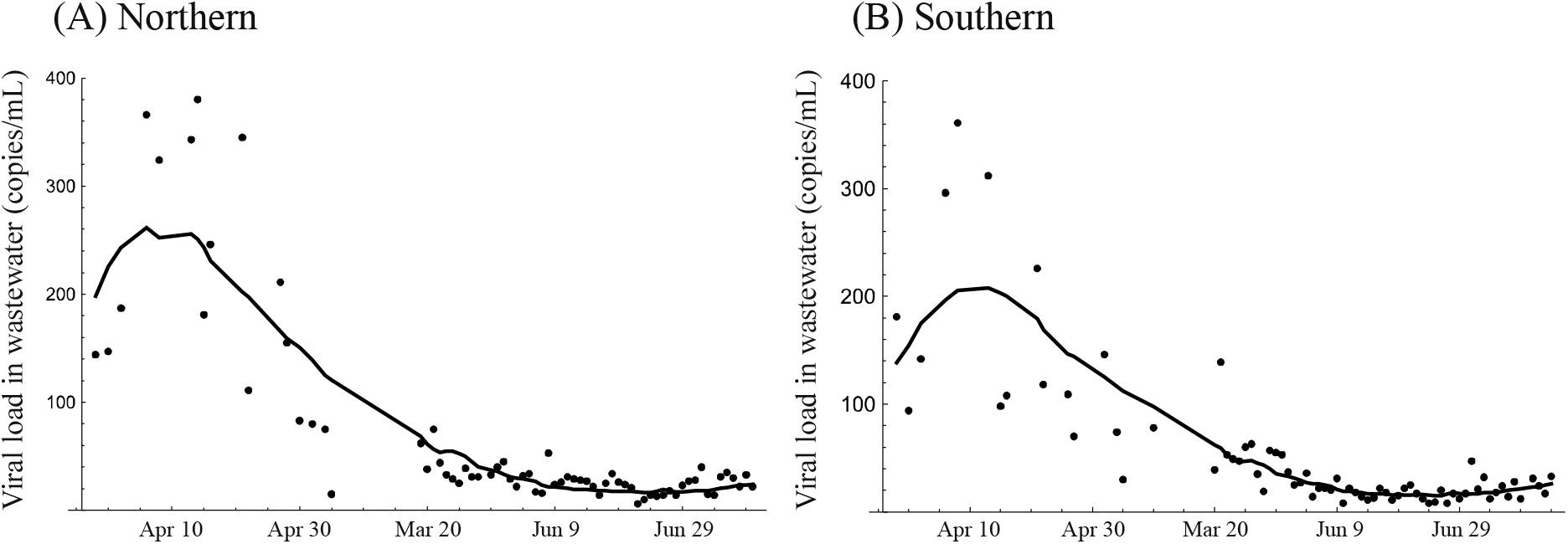
Model fittings to the viral load in wastewater sampled in the (A) northern and (B) southern areas. Solid lines show the model prediction and dots show the viral load in wastewater.

Our estimated length of the lag between the detection timing from wastewater and the reporting by passive surveillance suggested that the former can occur earlier than the latter, agreeing with a previous study (23). Compared to the incubation period of SARS-CoV-2 (24-26), our estimate of τ suggests that the virus is discharged from the body to sewers at early stages of infection. These results also suggest the potential success of early outbreak detection using wastewater. The estimated lag length was similar between both areas. The lag length was related to the natural history of the pathogen (i.e., the timing of virus discharge from body to sewer, and the timing of symptom onset, which triggers testing in passive surveillance) and testing capacity. These factors were likely similar between both areas because of geographical proximity.

The contribution of reported cases from the passive survey in age group 80+ yr to the viral load in wastewater was larger than that of 0–19 yr, a trend that was similar between both areas. This estimate suggests that the amount of virus shedding to wastewater among young individuals was smaller than that among elderly individuals. Furthermore, since the rate of asymptomatic infection among young individuals is higher than that among the elderly (4), the amount of virus shedding to wastewater among individuals with asymptomatic infection may be smaller than those with symptomatic infection. The number of confirmed cases from passive surveillance largely depends on the rate of symptomatic infection. If the amount of virus shedding to wastewater depends on the severity of symptoms, wastewater surveillance will also be affected by the rate of symptomatic infection.

The goodness of fit was better for the decreasing parts of the time series (Fig. 2). This might be simply because the beginning of the time series contained too much uncertainty, but another possible interpretation is that the estimability of parameters is ensured by an observation period that contains rich information on the clearance of viruses from infected individuals. In such a case, the difference in shedding kinetics among age groups determines the temporal changes in the viral load in wastewater. Although previous studies have investigated the duration of shedding or its time course with mainly symptomatic patient data (27-29), further studies on the determinants of the excretion characteristics of SARS-CoV-2 are needed.

## 4. Conclusion

The estimated lag between the time series of viral loads in wastewater and of reported cases in passive surveillance suggested the predictability of reported cases ten days later using viral loads in wastewater. Our modeling of the association between passive and wastewater surveillance showed that the contribution of a reported case in passive surveillance to the viral load in wastewater differed by age, emphasizing the importance of understanding age-dependent shedding kinetics. Further insights into the relationship between the amount of virus discharge to wastewater, age, and severity of symptoms would improve the predictive ability of WBE and make it more robust as a surveillance system.

## Data Availability

All data that support the findings of this study are openly available.

https://www.mwra.com/biobot/biobotdata.htm

https://www.mass.gov/info-details/archive-of-covid-19-cases-in-massachusetts

## Abbreviations

SARS-CoV-2: Severe acute respiratory syndrome coronavirus 2
WBE: wastewater-based epidemiology
PMMoV: pepper mild mottle virus

## Acknowledgment

This work was supported by JST, CREST (Grant Number JPMJCR20H1), JSPS KAKENHI (Grant Number 20J00793), Grants-in-Aid for Cross departmental Young Researcher Grants of Hokkaido University, Japan. The authors would like to thank Editage for the English language review.

## CRediT author contribution statement

Ryosuke Omori: Conceptualization, Methodology, Formal analysis, Writing - Original draft, Funding acquisition, Fuminari Miura: Methodology, Data curation, Writing - Original draft, Masaaki Kitajima: Writing - Original draft, Funding acquisition

